# Towards Personalized Edge-AI for Medicine: Efficient Neuromorphic Frameworks for Seizure Detection and Prediction

**DOI:** 10.1101/2025.09.22.25336341

**Authors:** Luis Fernando Herbozo Contreras, Leping Yu, Zhaojing Huang, Isabelle Aguilar, Armin Nikpour, Omid Kavehei

## Abstract

Epilepsy affects approximately 1% of global population, with 30-40% of cases resistant to conventional pharmacological treatments. Neurostimulation offers these patients an alternative by delivering targeted electrical stimulation to the brain. However, adaptive AI for these devices relies on external or cloud-based resources for model retraining and updating, which introduces significant challenges, including high false positive rates, latency issues, and privacy concerns. We present a neuromorphic framework for real-time seizure detection and prediction, directly implemented on a neuromorphic SOC with on-chip learning capabilities. By leveraging spiking neural networks and few-shot edge learning, our system enables continual, patient-specific adaptation without requiring data transmission or cloud connectivity. For a detection framework, a model pre-trained on the TUH dataset is deployed on the BrainChip Akida processor and personalized using long-term EPILEPSIAE recordings. For prediction, we demonstrate robust performance on both scalp and intracranial EEG from the CHB-MIT and Freiburg datasets using a leave-one-seizure-out strategy for edge training. Across datasets, our approach achieved commendable and superior performance to state-of-the-art neural networks in metrics as AUROC, Sensitivity, and False Positive Rates (FPR), while drastically reducing memory footprint through quantization and lowering energy consumption by orders of magnitude. This work establishes a foundation for next-generation personalized Edge-AI systems capable of adapting and learning directly at the edge.

## Introduction

Epilepsy is a chronic neurological disorder affecting more than 65 million people globally, characterized by recurrent seizures arising from aberrant electrical discharges in the brain. Clinical manifestations span from transient lapses of awareness to prolonged convulsive episodes, all of which impose profound burdens on the patient’s quality of life. Although advances in pharmacotherapy have improved outcomes, approximately 30–40% of individuals remain refractory to medication, highlighting the need for alternative interventions [1, 2]. Neurostimulation modalities such as the Responsive Neurostimulation System (RNS) have demonstrated efficacy in reducing seizure frequency in drug-resistant cohorts by sensing and sending electrical stimulation to the brain. Nevertheless, these systems exhibit several inherent limitations.

First, existing neurostimulation platforms rely on pre-trained detection models that often inadequately represent the considerable inter-patient heterogeneity in seizure morphology and dynamics, yielding elevated rates of both false positives and false negatives. Second, current AI-based personalization approaches generally perform model training and adaptation on external or cloud-based computing resources which leads to communication latency during acute seizure events and introduces potential privacy vulnerabilities [3]. Third, power and bandwidth constraints on implantable devices restrict intracranial electroencephalography (iEEG) telemetry to approximately one hour per day [4]. While extending this window of data could enhance model training, it would accelerate battery depletion and compromise device longevity.

Although model generalization measured via performance on distinct out-of-sample datasets is the prevailing metric for assessing adaptability to novel data, it does not necessarily predict performance on the continuously evolving biosignals of a single patient [5]. Accordingly, we advocate shifting the focus from population-level generalization to individual-level personalization, wherein a detection and prediction algorithm continuously refines itself to an individual’s unique neural signature at the edge. Personalized medicine, which aligns therapeutic strategies to an individual’s physiological profile, has demonstrated superior clinical utility across numerous domains [6, 7]. In epilepsy management, real-time, patient-specific adaptation of seizure detection and forecasting algorithms promises to revolutionize neurostimulation efficacy. However, realizing such personalization necessitates overcoming challenges in real-time signal processing, ultra-low-power operation, and resource-constrained computation [8]. To realize this shift from cloud-based AI to on-device intelligence, it is essential to review existing system-on-chip (SoC) solutions for seizure monitoring and their current limitations. Studies have demonstrated impressive advances in energy-efficient online training, achieving sensitivities (true positive rate) above 90%, specificities (true negative rate) exceeding 95–99%, and false-alarm rates below one event per hour while operating within the *µ*J-per-classification range [9–14]. These architectures are based on Convolutional Neural Networks (CNNs) [10], Support Vector Machine (SVM) and tree-based processors [9, 12], and unsupervised online classifiers [11] illustrate the maturity of low-power seizure detection hardware. Nevertheless, several common limitations remain:

- **Slow adaptation:** Training and retraining cycles typically occur over seconds or longer.
- **Limited adaptability:** Most designs are detection-only systems without continual personalization and evaluations are often confined to short-term datasets (i.e CHB-MIT) and small patient cohorts, limiting insights into long-term variability.
- **Frame-based processing:** Computation remains clock-driven, operating on fixed feature windows instead of leveraging sparse, event-driven nature of neural activity.
- **Incomplete reporting:** Performance metrics are primarily limited to sensitivity, specificity, and accuracy, with little emphasis on threshold-independent measures such as AUROC, reducing cross-study comparability.

These factors highlight the need for architectures capable of millisecond-scale continual learning, operation over long-term intracranial and scalp EEG recordings, and efficient event-based computation capabilities. Neuromorphic computing offers a compelling pathway to meet these demands by replicating the brain’s event-driven, energy-efficient processing paradigm. Compared to conventional digital architectures, neuromorphic systems can reduce computational energy consumption by several orders of magnitude [15], while supporting on-chip learning for continuous model adaptation without cloud reliance [4, 16]. Recent work has demonstrated online seizure detection [17] and the detection of High-Frequency Oscillations (HFO) as epilepsy-related biomarkers [18] using neuromorphic approaches. However, these implementations are restricted to inference and do not support continual learning. Such on-chip adaptability is essential to assimilate new data incrementally without catastrophic forgetting, ensuring sustained performance during physiological drift, whether due to electrode repositioning, evolution of seizures, or pharmacological effects [19, 20].

The key question motivating this work is whether it is possible to design a unified framework that bridges the gap between the state-of-the-art SoC’s and the adaptive, event-driven principles of neuromorphic computing for the next generation of neurostimulation devices. In this study, we present a personalized neuromorphic framework for on-device seizure detection and prediction that performs few-shot learning on a low-power hardware. This approach provides a scalable, privacy-preserving, and low-latency solution for adaptive neurostimulation. Our contributions include:

- Continual on-chip learning enables sustained performance improvement as new data arrive, reducing false-positive rates while enhancing sensitivity in both pipelines for 3 epilepsy datasets (2 long-term recording and 1 short-term recording).
- Rapid personalization is achieved with as little as three minutes of patient-specific seizure data (15-shot learning).
- On-device processing enables real-time adaptation and data privacy.
- Hardware-efficient training confined to final layer reduces computational cost.
- Energy and latency improve by 100 times compared with conventional architectures.
- Robustness in handling missing or corrupted EEG channels, simulating real-world scenarios of partial signal loss.

## Results

Fig. 1 illustrates our proposed edge-based personalized seizure detection and prediction framework. Unlike conventional epilepsy management methods that rely on cloud-or server-based AI, our approach envisions a future deployment scenario in which a neuromorphic system-on-chip could be integrated with existing closed-loop neurostimulation devices to enable on-device intelligence under stringent power constraints. The framework employs spike-encoded continuous Short-Time Fourier Transform (STFT) representations of EEG signals, processed through an offline-trained convolutional feature extractor on the BrainChip Akida [21], with only the final classifier adapted online to patient-specific data. For seizure detection, a CNN trained on the TUH-EEG corpus is quantized, converted to a SNN, and edge-refined using the EPILEPSIAE dataset. For seizure prediction, a similar pipeline is applied to the Freiburg iEEG and CHB-MIT dataset, with models pre-trained on a GPU and then updated at the edge, ensuring adaptability and independence from cloud connectivity.

**Fig. 1.**
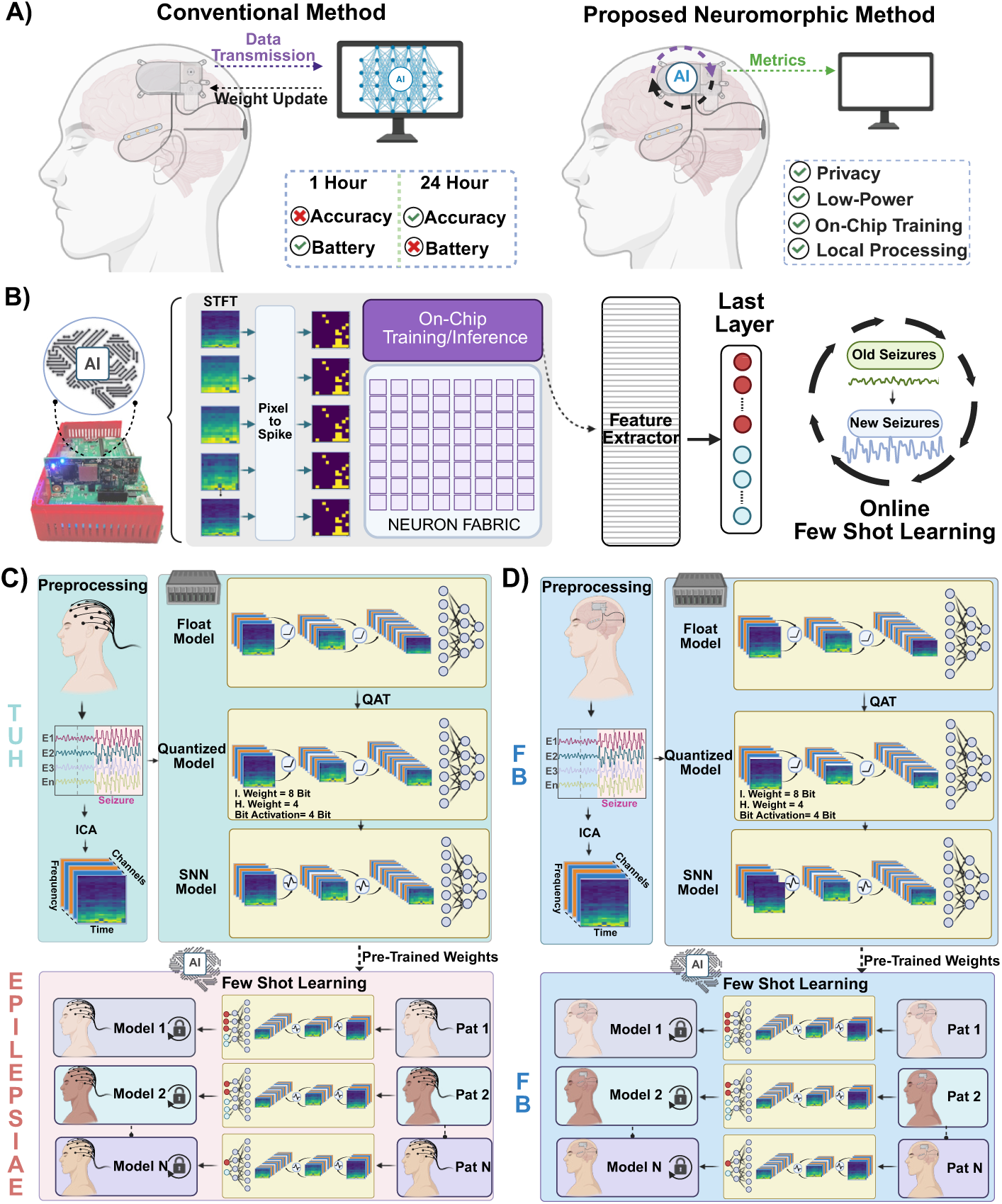
Edge-based personalized seizure detection and prediction frameworks. Conventional Meth­ods for treating epilepsy rely on AI outside the device, while our proposed method envisions an efficient neuromorphic-AI system-on-chip closed-loop neurostimulation device without compromis­ing power consumption (A). Continuous Short-Time Fourier Transform (STFT) of EEG images is spike-encoded and passed through a frozen, pre-trained convolutional feature extractor at the edge. Only the final classifier layer is updated on-chip via online few-shot learning, reducing compute and memory overheads (B). For the detection framework, a CNN is trained on a large-scale dataset (TUH), quantized to mixed precision and converted to a Spiking Neural network (SNN), and edge-adapted with patient-specific data from a long-term recording dataset (EPILEPSIAE) for patient-specific (C). For the prediction pipeline, the same methodology is applied to the Freiburg and CHB-MIT datasets. Because of limited patient numbers, individualized models are pre-trained rather than a global model and subsequently refined on previously unseen patient data at the edge, yielding an adaptable cloud-independent approach (D).

**Table 1.** Summary of EEG datasets used in this study.

| Dataset<sup>◊</sup> | Type | Rate (Hz) | No. of Patients | No. of Channels | No. of Seizures | Interictal time(h) | Task | Edge Train | Temporal Duration |
| --- | --- | --- | --- | --- | --- | --- | --- | --- | --- |
| TUH | Scalp | 250 | 642 | 19 | 3050 | 859.7 | Detection | No | Short-Term |
| EPILEPSIAE | Scalp | 250 | 30 | 19 | 276 | 4598.2 | Detection | Yes | Long-Term |
| CHB-MIT | Scalp | 256 | 13 | 22 | 64 | 209 | Prediction | Yes | Short-Term |
| FB | Intracranial | 256 | 13 | 6 | 59 | 311.4 | Prediction | Yes | Long-Term |

◊ Temple University EEG Corpus (TUH), Boston Children’s Hospital–MIT (CHB-MIT) and Freiburg Hospital (FB).

### Feature Extraction

An essential preliminary step for enabling edge learning is to ensure that the backbone models capture physiologically meaningful neural dynamics. Before STFT feature extraction, ICA-based artifact rejection was applied to the scalp EEG recordings. Independent components strongly correlated with the frontal FP1 and FP2 channels were identified as ocular-related components and removed, as detailed in the Methods section.. We first demonstrate the STFT representations of a dataset for the non-seizure and seizure classes. We then extracted saliency maps from the trained model using Gradient-weighted Class Activation Mapping (Grad-CAM) for both classes, which reveal consistent patterns of model attention, with a pronounced emphasis on lower frequency regions. Results are shown in Fig. 2(B). This observation aligns with prior findings that low-frequency activity and slow-wave components are key biomarkers associated with seizures in scalp EEG recordings [22].

**Fig. 2.**
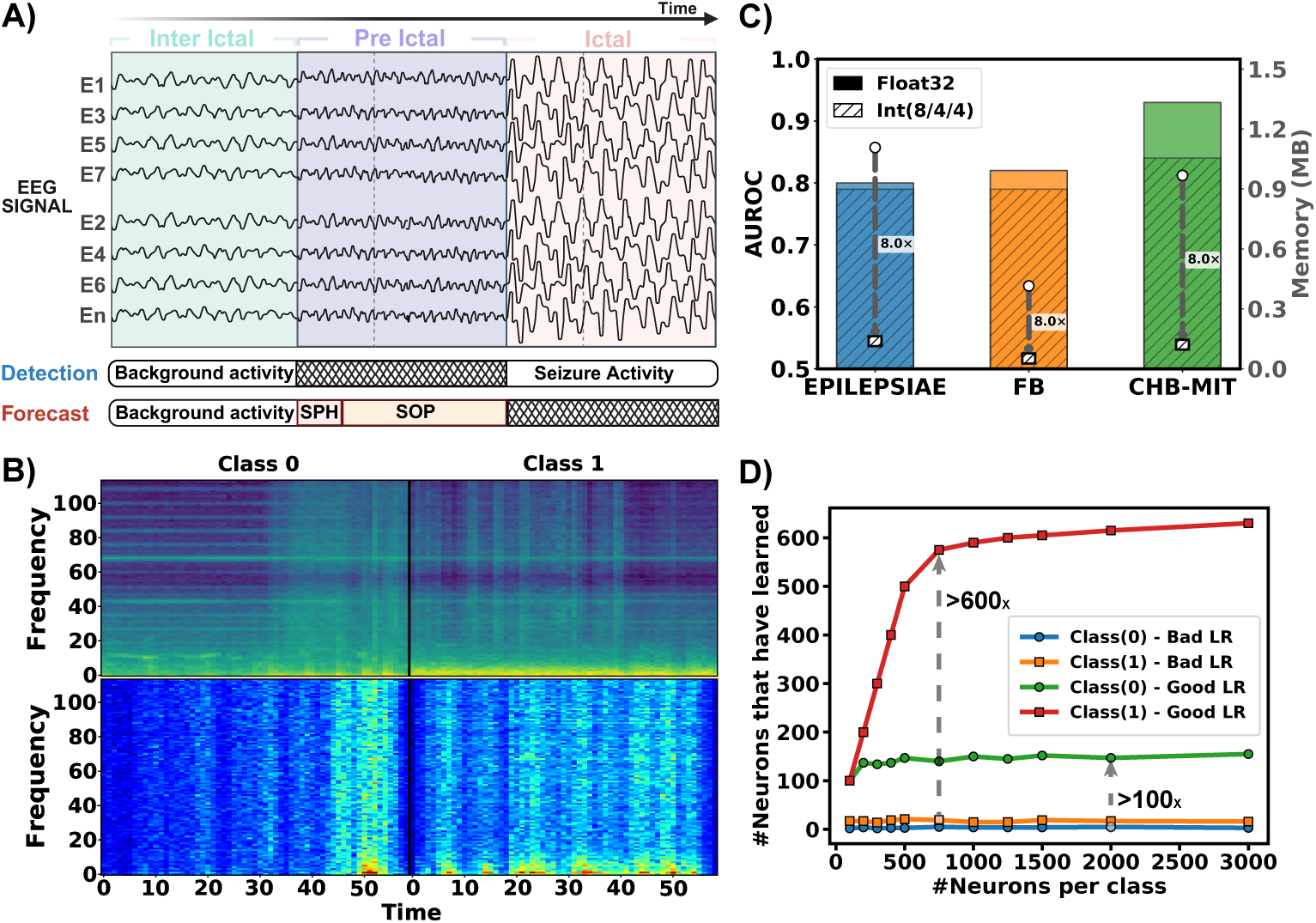
Results for the Initial Training and Prediction Stage on GPU. When analyzing seizure events, three phases are particularly critical: interictal, preictal, and ictal. These phases form the basis of two distinct tasks: Detection (discriminating between interictal and ictal states) and forecasting (discriminating between interictal and preictal states) (A). We applied STFT to the EEG data to obtain representations of Class 0, denoting interictal (non-seizure) activity, and Class 1, denoting ictal (seizure) activity. The Grad-CAM maps, in which warmer colors denote higher importance scores for the corresponding class, revealed distinct class-specific activation patterns predominantly at lower frequencies (B). Across all datasets, our models achieved commendable AUROC scores under floating-point precision, with only slight reductions after quantization, while still yielding up to 8 times reduction in memory (C). During quantization, lowering the learning rate relative to the initial training phase proved essential, as it allowed more effective consolidation of previously learned neuronal activations before transitioning to edge learning. Notably, proper initialization with an appropriately tuned learning rate boosted per class neuron activations from 100 to over 600 times(D).

### Pre-Post Quantization

In the initial stage, we evaluated model performance on the TUH, FB, and CHB-MIT datasets using AUROC, recall, and precision. Model weights with validation scores exceeding or achieving 0.80 (0.80, 0.82, and 0.93, respectively) were selected for weight transfer. Following quantization, performance declined only marginally for the TUH and FB datasets (1% and 3%, respectively), whereas the CHB-MIT dataset exhibited a more pronounced reduction of 9%. Despite this, memory requirements were reduced by approximately 8 times across all datasets as an outcome of the adopted quantization scheme. Comparative AUROC results for both non-quantized and quantized models, alongside memory consumption (MB), are presented in Fig. 2(C).

### Quantization Learning Rate effects on Neuron Activations

We set up the learning rate to be 10 times lower than the original learning rate that was used to compile in the training GPU phase. Through our observations, this has the biggest impact on the edge deployability. A bad learning rate (to be similar to the stage 1 of training) leads to fewer neurons been learned per class, as observed in Fig. 2(D) for non-seizure and seizure classes, respectively. A neuron is considered “learned” when its learning threshold becomes positive during edge training, indicating that it represents a feature pattern associated with a given class.. We observed as the number we input, with a good initialization of LR there was neurons learned for both classes. As well, this behavior also gives insights that when performance improvements became marginal beyond a certain point, the number of neurons was capped. In this way, resources were allocated efficiently, reducing memory usage on the BrainChip Akida hardware without compromising performance. For instance, as illustrated in Fig. 2(D), increasing the final-layer beyond 1000 neurons led to only a small increase in the number of neurons allocated to each class, indicating saturation. We therefore capped the final layer at 1000 neurons to avoid memory consumption.

### Detection Out-Of-Sample Framework

Using the transferred weights, we further trained the model on the EPILEPSIAE dataset directly at the edge. To provide a comprehensive assessment of performance, we introduce a Figure of Merit (FoM) that combines multiple evaluation metrics into a single score. The FoM is defined as:

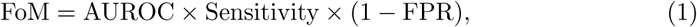

where (1-FPR) is choosen to penalizes false alarms. Sensitivity reflects a True Positive Rate. This formulation enables a balanced evaluation across competing objectives of accuracy, reliability, and robustness. Fig. 4 presents the FoM (Eq. 1) as a function of the number of shots, with corresponding latency measurements plotted in parallel in all datasets. As the number of shots increases, all performance metrics exhibit consistent improvement. By the 15th shot, the AUROC, sensitivity, and FPR attained values of 0.85, 0.87, and 0.20, respectively. These personalized outcomes were subsequently compared against a generalization baseline, which reflects performance under inference-only conditions without further adaptation and with floating precision (float32). The personalized models demonstrated superior performance, yielding relative gains of 2% in AUROC, 4% in sensitivity, and a 6% reduction in FPR. These findings demonstrate that continuous, on-device learning with patient-specific data is both beneficial and critical for achieving effective, personalized seizure detection. The dataset holds particular significance as it comprises long-term continuous recordings in which seizure events constitute less than 1% of the total duration, with interictal segments comprising nearly the entirety of the dataset. Results are shown in Fig. 3(A). Detailed per-patient results are provided in Supplementary Table S1. Few-shot learning results across all patients are shown in Supplementary Fig. S2.

**Fig. 3.**
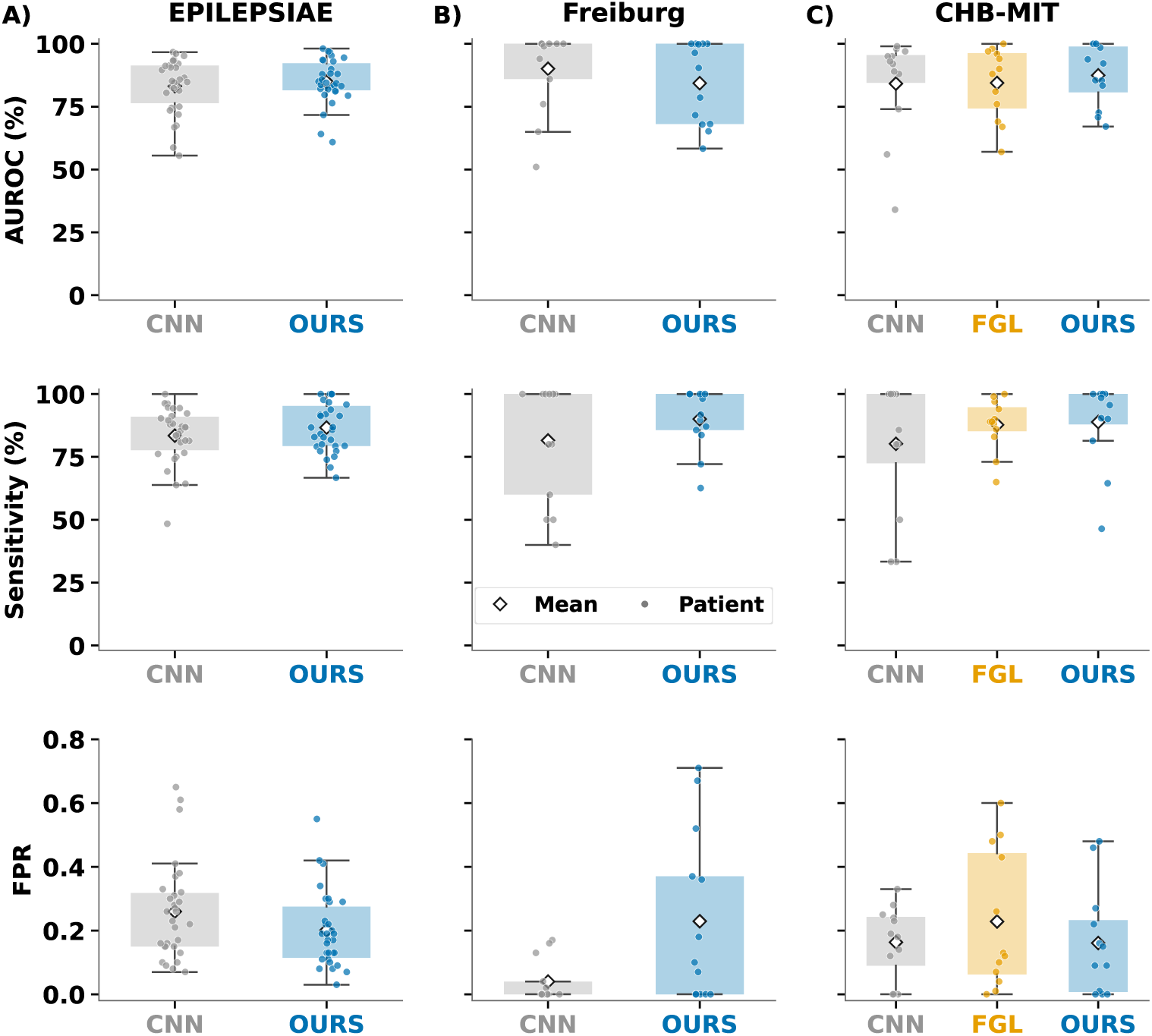
Performance comparison across all datasets. Distributions of AUROC, sensitivity, and false positive rate (FPR) are presented for EPILEPSIAE (A), FB (B), and CHB-MIT (C) against an established and recent state-of-the-art baseline. It is worth noting that our evaluations were conducted under more resource-constrained conditions than those used by others models [23, 24].

**Fig. 4.**
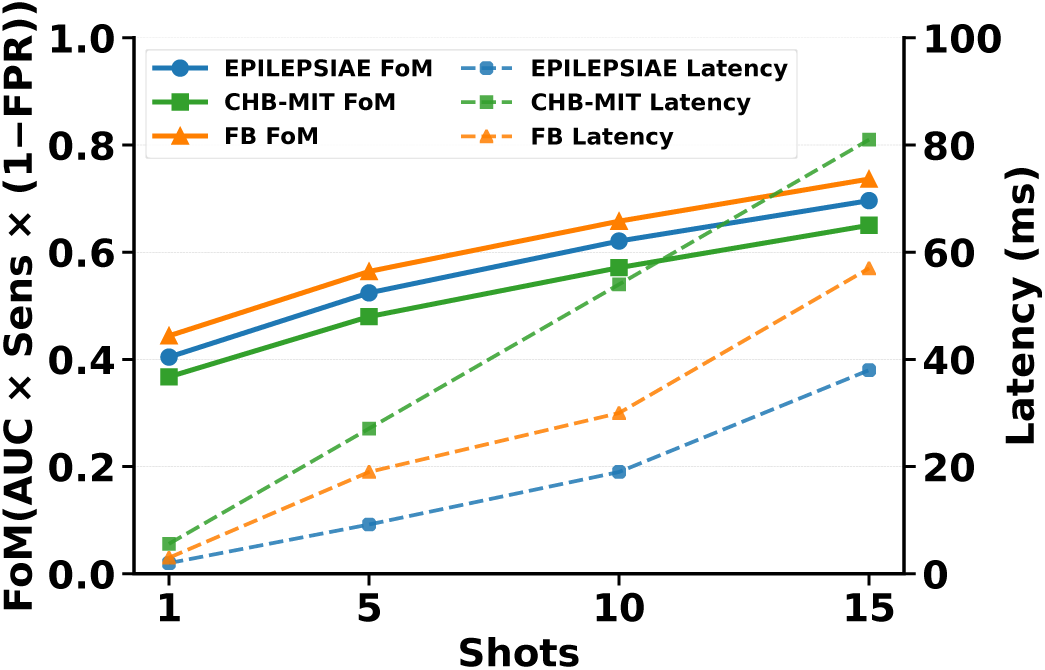
Figure of Merit (FoM) and latency vs number of shots for the three datasets. FoM is defined as *AUROC × Sensitivity ×* (1 *− FPR*). Higher shot counts yield consistent FoM gains, while latency rises marginally, indicating improved performance without meaningful computational cost.

### Prediction Framework

For the prediction task, we employed two datasets to compare different baselines: the scalp EEG dataset (CHB-MIT) and the iEEG dataset (FB). The inclusion of the FB dataset also aligns with the vision of self-sustaining AI at the edge for future implantable devices. The experimental setup differed from the TUH-EPILEPSIAE framework, where models were first trained on TUH using a high-performance GPU and subsequently adapted at the edge on the EPILEPSIAE dataset. In contrast, for both CHB-MIT and FB, models were initially trained on a GPU, and then a leave-one-seizure-out approach was used for edge learning. This strategy reserved one seizure per patient for on-chip training while leaving sufficient samples for post-training evaluation, ensuring robust assessment of edge adaptation across both datasets. For preprocessing, we followed the same pipelines in two studies [23, 24].

For the CHB-MIT dataset, the floating-point model achieved an AUROC greater than 90% during GPU training, followed by an approximately 9% reduction after quantization (Fig. 2(C)). Using the pre-trained individual weights, we subsequently trained patient-specific models at the edge. As the number of shots increased up to 15, performance stabilized with AUROC, sensitivity, and FPR reaching 0.87, 0.89, and 0.16, respectively (see Supplementary Fig S1). To benchmark our approach, we compared it with an established CNN reference baseline and a recent state-of-the-art methodology. The first baseline was a widely cited CNN architecture for seizure prediction [23], trained in full-precision and evaluated only at inference. This model achieved AUROC and sensitivity scores of 0.84 and 0.80, respectively, whereas our method improved upon these results by 3% in AUROC and 9% in sensitivity. The second baseline was the recently proposed Future-Guided Learning (FGL) framework [24], which integrates detection models to support prediction training and has demonstrated competitive performance against established benchmarks. Notably, our approach outperformed FGL across all metrics, achieving relative improvements of 3% in AUROC, 1% in sensitivity, and 7% in FPR. Results are shown in Fig. 3(C). Detailed per-patient results are provided in Supplementary Table S2. Results of few-shot learning across all patients are shown in Supplementary Fig. S3(B).

For the FB dataset, results show a similar trend to the previous model, with smaller performance gains but greater stability in metrics across shots (see Supplementary Fig. S1). This limitation can be attributed in part to the BrainChip Akida architecture’s requirement to use odd kernel sizes, which led us to mask one channel to conform to the hardware constraints. As discussed earlier, the first convolutional kernel spans the available channels, and given that the FB dataset includes only six intracranial channels, this restriction likely contributed to the diminished improvement. Despite these constraints, our edge-trained model achieved AUROC, sensitivity, and FPR values of 0.84, 0.90, and 0.23, respectively, at 15 shots. Our sensitivity increased by nearly 10% compared with the established CNN reference baseline for seizure prediction. However, our false-positive rate remained higher (0.23 vs 0.06), and the AUROC was approximately 4–5% lower than the CNN baseline. It is important to note, though, that the baseline CNN was trained in full precision (float32) and utilized all available intracranial channels, whereas our approach was implemented under the quantization and one less channel constraints of the Akida neuromorphic hardware. Results are demonstrated in Fig. 3(B). Detailed per-patient results are provided in Supplementary Table S3. Few-shot learning results across all patients are shown in Supplementary Fig. S3(A).

### Power, Memory and Latency Analysis

The power consumption and inference efficiency of the Akida system were systematically evaluated using the MetaTF power profiling API during the inference stage [25]. The floor power, representing the dynamic baseline power consumption of the device while idle, was measured at 900.5 mW. During active inference, the system achieved an average throughput of 70.41 frames per second (fps). Each frame corresponds to the STFT representation of a 12-second EEG window. The inference latency per frame was 14.2 ms, which equates to approximately 1.18 ms per second of raw EEG data. The energy consumption per inference was measured at 12.86 mJ per frame of a 12-second EEG window, which is equivalent to 1.07 mJ per 1 second of EEG data, reflecting the low-power profile of the neuromorphic hardware. These results demonstrate the suitability of the Akida platform for real-time, energy-efficient processing of EEG data in edge-based seizure detection and prediction applications. Results can be found in detail in Fig. 5, where we compare running the CNN backbone in a conventional CPU architecture and our approach in the Akida to see the latency and energy per inference. We demonstrate that our neuromorphic approach enables low power consumption by two to three orders of magnitude.

**Fig. 5.**
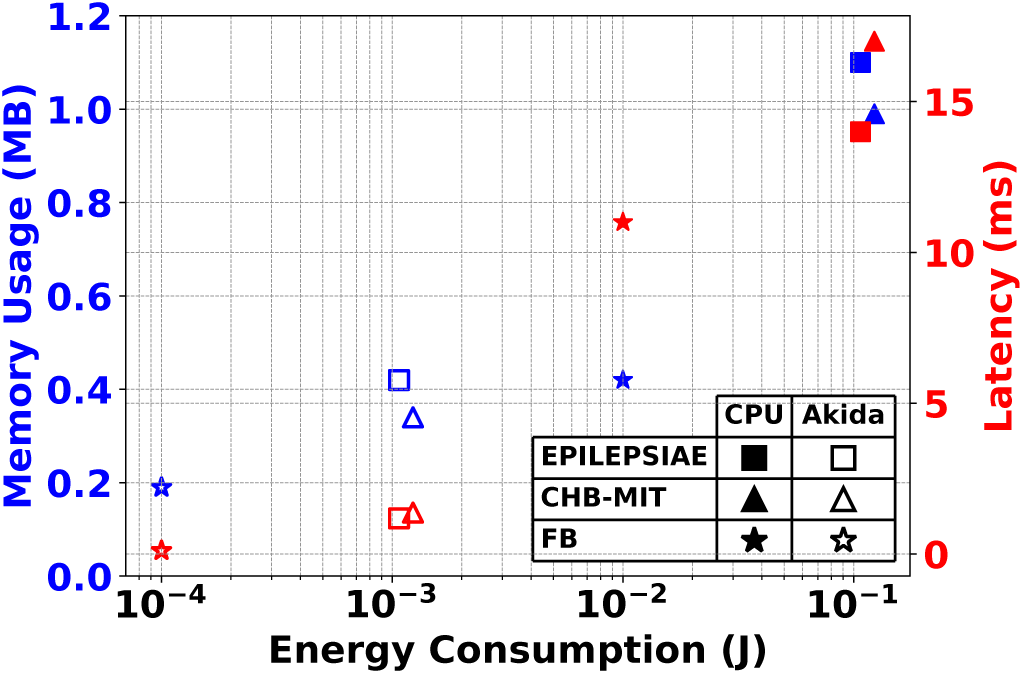
Pareto plot of energy consumption versus Memory Usage and Latency for three datasets (EPILEPSIAE, CHB-MIT, and FB). Filled markers represent the results for metrics in CPU, while non-filled markers represent those at the Edge with Akida.

### Robustness

A critical validation in this study was to examine the robustness of our model in the spiking domain under conditions of reduced input information. We evaluated this on the EPILEPSIAE and CHB-MIT datasets, each comprising 19 EEG channels, by setting 20%, 40%, and 60% of the input channels to zero. At each blackout level, channels were sampled uniformly at random without replacement and were not constrained to consecutive blocks. Three independently generated channel masks, obtained using distinct predetermined random seeds, were evaluated at each level, and the resulting performance metrics were averaged. As illustrated in Fig. 6, the CHB-MIT dataset demonstrated stable performance despite increasing channel loss and further benefited from a higher number of shots. In contrast, the EPILEPSIAE dataset showed a more pronounced degradation as blackout levels increased. Nevertheless, resilience improved considerably with additional shots. We attribute this effect to the long-term nature of EPILEPSIAE recordings, which represent a more realistic and challenging benchmark compared with CHB-MIT. Overall, our model achieved strong performance across both datasets, highlighting its robustness under conditions of channel dropout.

**Fig. 6.**
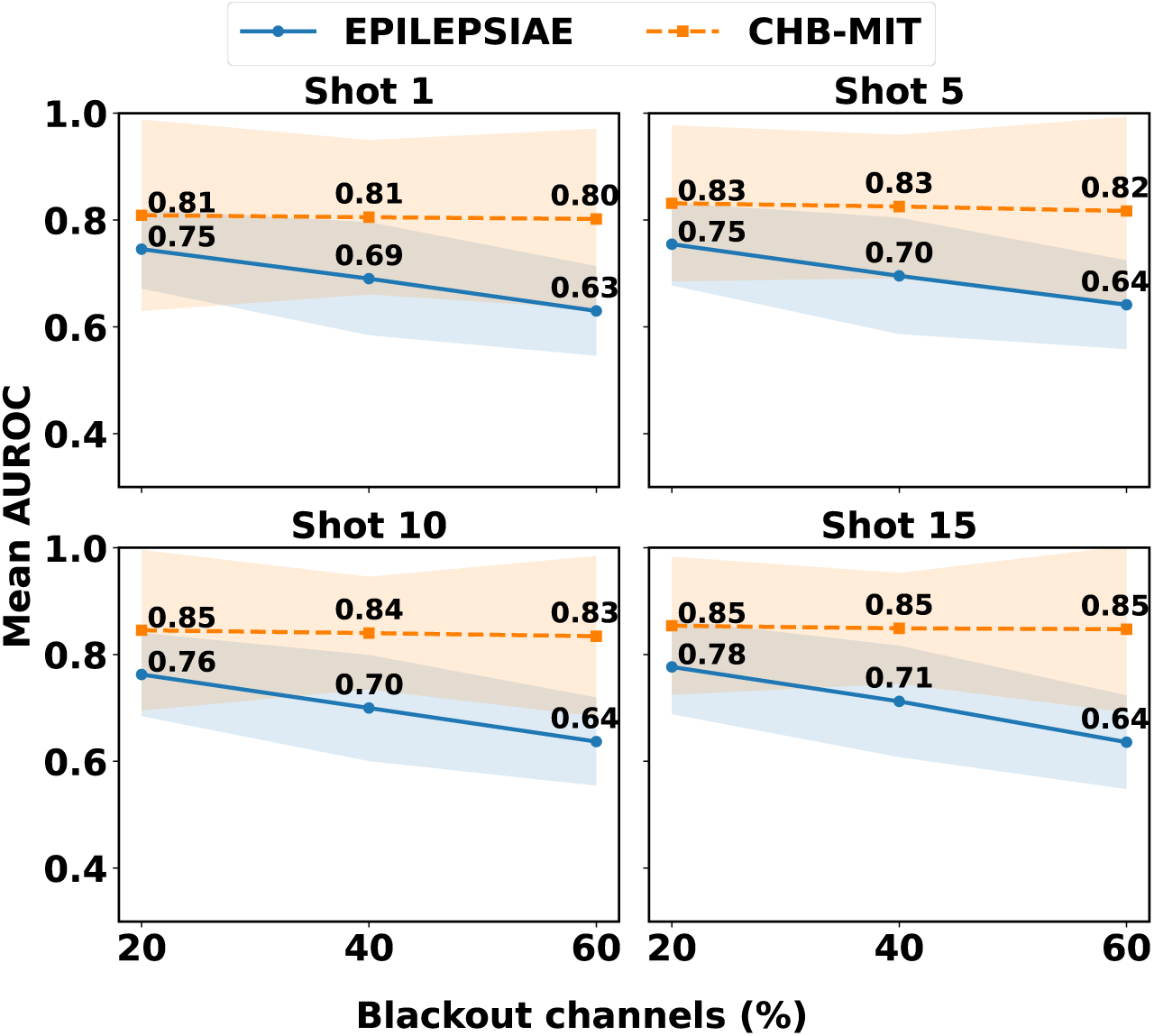
Robustness test of our approach for seizure detection and prediction across datasets under progressive channel blackout with three repetitions. Performance is shown for the EPILEPSIAE dataset (blue, long-term clinical data) and the CHB-MIT dataset (orange) across different training shots (1, 5, 10, 15) as increasing proportions of EEG channels were set to zero, highlighting resilience to missing-channel conditions.

### Complexity: Training and Inference

Our analysis of computational complexity highlights the efficiency of the proposed Akida-based edge learning framework compared to conventional deep learning approaches. Traditional CNNs and RNNs exhibit high training costs due to their dependence on large parameter updates and, in the case of recurrent models, sequential operations that limit parallelization[26]. Similarly, SNNs trained with surrogate gradients require costly backpropagation through time, while even local learning rules still scale linearly with sequence length [27]. SVMs, though non-sequential at inference, demand prohibitively expensive training with cubic dependence on the number of samples [9, 10]. By contrast, our proposed edge learning approach reduces training as a cluster with linear complexity in the feature dimension, while the inference scales with the number of neurons and the feature extractor. Crucially, this approach enables efficient, low-latency operation. These findings demonstrate that EDGE-AI on neuromorphic hardware offers a favorable trade-off between performance and computational cost, positioning neuromorphic as a viable solution for real-time, resource-constrained biomedical applications. The complexity formulas can be found in Table 2. We benchmark our approach against representative state-of-the-art seizure detection/prediction models whose architectures and hyperparameters are documented in the literature and evaluated on at least three EEG datasets. In Fig. 7, we demonstrate that our approach is computationally efficient for both training and inference as it occupies the lower-left region of the plot.

**Fig. 7.**
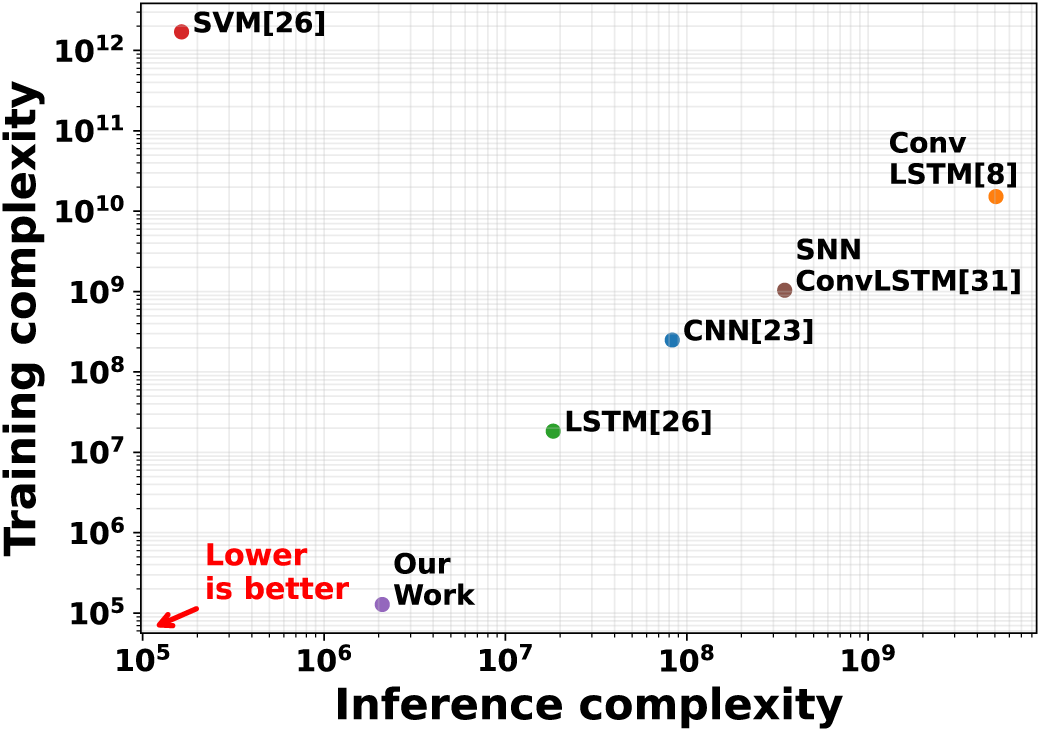
Log–log scatter plot of training vs inference computational complexity for representative EEG seizure detection/prediction models. Each point reflects hyperparameters reported in prior work. Conventional method of training these networks are BP (CNN) and BPTT (LSTM, SNNConvLSTM, ConvLSTM). For the proposed method, training complexity represents only on-device adaptation of the final classification layer; offline pretraining and QAT are excluded. For inference complexity, the feature extractor is taken into account as a SNNConv.

**Table 2.** Training complexity per model.

| Algorithm | Training Complexity<sup>⊗</sup> |
| --- | --- |
| CNN<sup>⊙</sup> | $\mathcal{O}\left(\sum_{\ell=1}^L C_{\text{in}}^{(\ell)} C_{\text{out}}^{(\ell)} K_{\ell}^2 H_{\ell} W_{\ell}\right)$ |
| LSTM<sup>†</sup> | $\mathcal{O}\left(\sum_{\ell=1}^L T \cdot 4 \left( (M^{(\ell)})^2 + M^{(\ell)} D^{(\ell)} \right)\right)$ |
| ConvLSTM<sup>†</sup> | $\mathcal{O}\left(\sum_{\ell=1}^L T \cdot 4 \left[ C_{\text{in}}^{(\ell)} C_{\text{out}}^{(\ell)} K_{\ell}^2 H_s W_s + (C_{\text{out}}^{(\ell)})^2 K_{\ell}^2 H_s W_s \right]\right)$ |
| SNN<sup>†</sup> | $\mathcal{O}\left(\sum_{\ell=1}^L T \cdot \text{cost}_{\text{layer}}^{(\ell)}\right)$ |
| SNNConvLSTM<sup>†</sup> | $\mathcal{O}\left(\sum_{\ell=1}^L T \cdot 4 \left[ Fr_{x,\ell} C_{\text{in}}^{(\ell)} C_{\text{out}}^{(\ell)} K_{\ell}^2 H_s W_s + Fr_{h,\ell} (C_{\text{out}}^{(\ell)})^2 K_{\ell}^2 H_s W_s \right]\right)$ |
| SVM | $\mathcal{O}(n^2 m + n^3)$ |
| **Proposed** | $\mathcal{O}(n \cdot U)$ |

<sup>⊙</sup> Trained with Backpropagation (BP).
<sup>†</sup> Trained with Backpropagation Through Time (BPTT).
<sup>⊗</sup> L:Layer; H:Height; W:Width; K:Kernel; C:Filter; T:Sequence length; M:Hidden size; D:Input Size; n:Neurons; m:Feature dim.; $Fr_{in}, Fr_{out}$ : Spike Rate; U:Input Spikes.

## Discussion

This study presents a novel framework for seizure detection and prediction directly on the edge, enabling continual learning with lower computational complexity, reduced energy consumption, and fully unsupervised adaptation while processing streaming EEG data. These characteristics make the framework particularly suitable for long-term neuromorphic neuromodulation, where embedded intelligence must operate reliably, efficiently, and with minimal dependence on cloud resources. While the results demonstrate strong potential, several opportunities remain for further improvement.

- In this study, we do not consider different structures of different neural networks, which can lead to better results. Nonetheless, by using a simple architecture, we are able to see the increase in performance, compared to more complex models[8, 23].
- BrainChip Akida only allows certain models and layers to run at the Edge for training. It can be more beneficial to incorporate more advanced algorithms that can run on the edge [27–31].
- Different encoding mechanism where not explored in this study. Studies have shown that different techniques in processing the EEG data into spikes is fundamental to capture temporal dynamics on seizure data [32] and also a reduction in power consumption and continual learning [33].

Future work will focus on designing architectures capable of better capturing temporal and spatial dependencies, as shown in recent studies [34], thereby achieving higher performance without increasing memory or computational requirements.

Overall, this work provides a promising foundation for next-generation AI-driven electroceuticals.

## Methods

### Datasets

For seizure detection, we utilized the TUH-EEG Corpus and the European EPILEPSIAE dataset. The TUH-EEG Corpus, featuring diverse recording conditions across 592 training patients and 50 validation sessions, provided a robust foundation for initial model development. To address real-world clinical scenarios, we employed the EPILEPSIAE dataset (30 patients) for personalized modeling via few-shot transfer learning. The dataset exhibits a significant class imbalance, characterized by a high ratio of interictal to ictal segments, serving to evaluate the adaptive performance of our algorithms during on-device personalization. For seizure prediction, the CHB-MIT (scalp EEG) and FB (iEEG) datasets were employed. From CHB-MIT, we selected 13 patients who met clinical relevance thresholds (fewer than 10 seizures per day and at least three leading seizures), treating clusters within 30 minutes as single events [35]. The FB dataset consists of recordings from 21 patients with refractory epilepsy, of which 13 were included in this study due to data availability constraints. Six channels were selected: three from epileptogenic regions and three from non-epileptogenic areas. The dataset includes at least 50 minutes of pre-ictal data and 24 hours of interictal recordings per patient, sampled at 256Hz. Further details are summarized in Table 1.

### Pre-Processing

To address challenges inherent in raw EEG signals, we employed two preprocessing techniques: Independent Component Analysis (ICA) and the Short-Time Fourier Transform (STFT). EEG recordings were first segmented into 12-second intervals, after which ICA was applied to perform Blind Source Separation (BSS), decomposing the signal into 19 statistically independent components. This process is represented in Eq. (2),

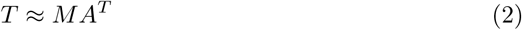

where *T* denotes the EEG signal matrix, *M* captures the temporal dynamics, and *A* contains the spatial weights corresponding to the topographic maps. To identify components related to ocular artifacts, we computed the Pearson correlation between each independent source and the ‘FP1’ and ‘FP2’ EEG channels. Components exhibiting strong correlations with eye movements were removed, yielding artifact-reduced EEG signals. Following this cleaning step, we applied the STFT with a 1-second window (250 samples) and 50% overlap, while also discarding the DC component. This transformation resulted in data shaped as (*N ×* 23 *×* 125), where *N* is the number of electrodes, 23 corresponds to the time steps, and 125 represents the frequency bins.

### Implementation Details

This study addresses two distinct temporal tasks: detection and prediction. As illustrated in Fig. 2(A), we categorize EEG data into inter-ictal, pre-ictal, and ictal states. Detection focuses on real-time classification between inter-ictal (background) and ictal (seizure) activity. In contrast, prediction aims to identify the pre-ictal state, specifically defining the Seizure Prediction Horizon (SPH) and the Seizure Onset Period (SOP). A prediction is considered correct if the seizure onset occurs after the SPH and within the SOP. Conversely, a false alarm arises when the system triggers a positive prediction but no seizure occurs during the SOP.

We pre-trained a compact CNN on the TUH dataset (200 epochs, Adam optimizer, MSE loss, *LR* = 0.005). The architecture consists of three convolutional blocks with increasing filter sizes (16, 32, 64), each followed by batch normalization, ReLU activation, and max-pooling. The first convolution spans all *N* EEG channels to capture spatial frequency dependencies [23]. Features are flattened and passed through a 128-unit dense layer with 0.5 dropout prior to the final classifier.

As Akida requires odd-dimensional kernels, one input channel was excluded from both the CHB-MIT and FB datasets to ensure hardware compatibility. For each dataset, Channel 0 was selected following visual comparison of the temporal and spectral representations of all available channels, as its feature patterns appeared distinct from those of the remaining channels.

To enable deployment on the Akida processor, Quantization-Aware Training (QAT) was applied using an 8-4-4-1 precision scheme (8-bit inputs, 4-bit weights and activations, and 1-bit output). The binary output activation enables Akida’s integer-only on-chip learning mechanism. QAT was performed using Adam optimizer with a learning rate of 5 *×* 10^−4^, *β*_1_ = 0.9, *β*_2_ = 0.999, and *ɛ* = 10^−8^. MSE was used as the loss function. Early stopping monitored the validation AUROC with a patience of 5 epochs. Model weights corresponding to the highest validation AUROC were retained. The trained quantized ANN was then converted into an SNN by replacing ReLU activations with spiking dynamics and transferring the quantized weights. Input encoding was performed using Rank Order Coding (ROC), in which information is conveyed by the relative order and location of events rather than by firing rates.

### Edge Learning Learning

Following SNN mapping, we performed edge learning in Akida by enabling an efficient on-chip adaptation through a two-phase process. First, a pre-trained, quantized backbone is deployed with frozen weights to preserve feature representations. Second, only the final classification layer remains trainable, serving as a clustering mechanism with the neuron count as a tunable parameter. To optimize this, we reserved 10–20% of the training data as an estimation set to analyze activation behavior and calibrate neuron allocation. Neuron counts between 100 and 3000 were assessed to optimize the accuracy-memory trade-off. Akida enables few-shot adaptation to new seizure patterns using minimally labeled data (1 shot = 12s). By fine-tuning only the final layer on the device, we achieved individual patient seizure characteristics without cloud dependency or full network retraining (Fig. 1C-D).

### Performance Metrics

The datasets used for model training and evaluation includes labeled data for 12-second time window, as previously used in papers as a baseline reference [16, 26, 31, 34, 36]. A seizure is identified when the model accurately predicts an ictal event (class 1) based on the ground truth, while non-seizure events are labeled as class 0. Seizure datasets are often imbalanced, with far more non-seizure data points, since seizures are rare and brief. Over a day, seizure occurrences represent only a small fraction of the data within a 12-second window. Traditional metrics (e.g. accuracy) are misleading as they tend to favor the majority class and do not effectively measure the model’s ability to detect seizures, which is the primary goal. For this reason, we rely on the Area Under the Receiver Operating Characteristic Curve (AUC-ROC), which evaluates both sensitivity and specificity in a threshold-independent manner. We also provided additional metrics, including Sensitivity and False Positive Rate (FPR).

## Declarations

### Data Availability

The TUH dataset can be freely obtained [here](https://isip.piconepress.com/projects/tuh_eeg/). Access to the EPILEPSIAE dataset requires payment and is accessible [here](https://www.epilepsy-database.eu/). The FB data used in this study was previously available openly via this [link](https://epilepsy.uni-freiburg.de/freiburg-seizure-prediction-project/eeg-database), but it seems it no longer accepts registration. The Children’s Hospital Boston dataset is publicly available [here](https://physionet.org/content/chbmit/1.0.0).

### Code Availability

All source code will be made publicly available upon acceptance of this manuscript.

## Acknowledgements

Luis Fernando Herbozo Contreras would like to acknowledge the partial support of the Faculty of Engineering Research Scholarship provided by The University of Sydney. Zhaojing Huang would like to acknowledge the support of the Research Training Program (RTP) provided by the Australian Government. The research is supported by the Australian Research Council under Project DP230100019.

## Author Contribution

LFHC developed the methodology, conducted experiments, prepared the figures, and wrote the draft manuscript. LY, ZH, and IA contributed to the drafting of the manuscript, the review of results, and the revision of the methodology design. AN and OK supervised experiments, manuscript preparation, reviewed methodology design and development, and reviewed results and manuscript drafts. OK developed the concept, framework conceptualization and served as the principal supervisor. All authors read and approved the final manuscript.

## Supplementary Information

### Supplementary Figures

**Figure S1:**
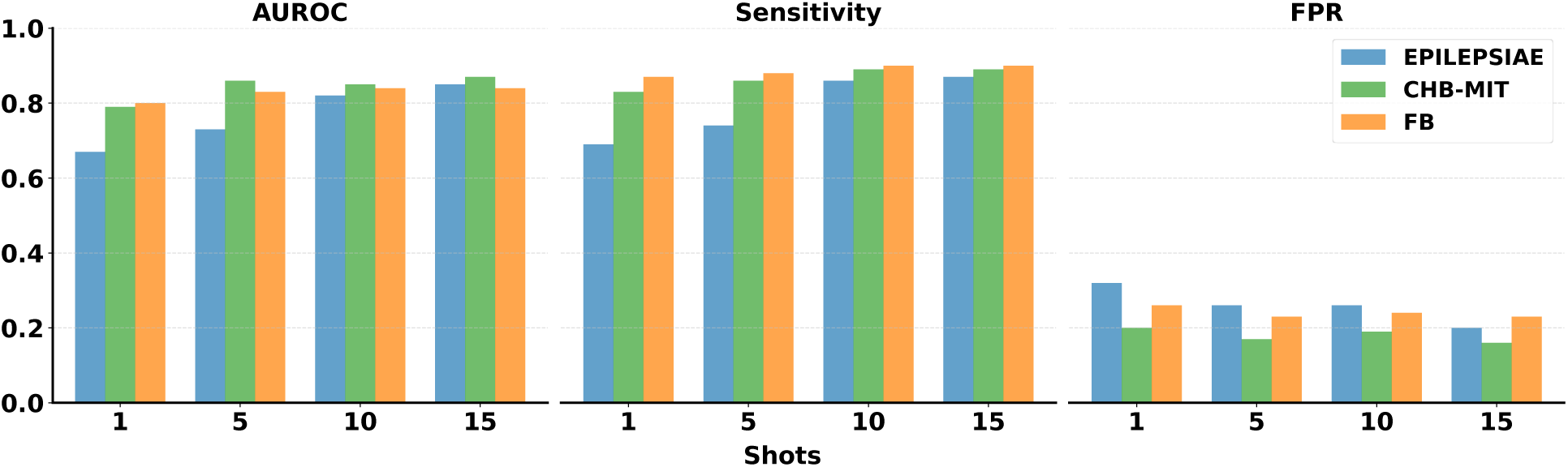
Average AUROC, FPR, and Sensitivity across all patients and datasets (EPILEP-SIAE, CHB-MIT, Freiburg)

**Figure S2:**
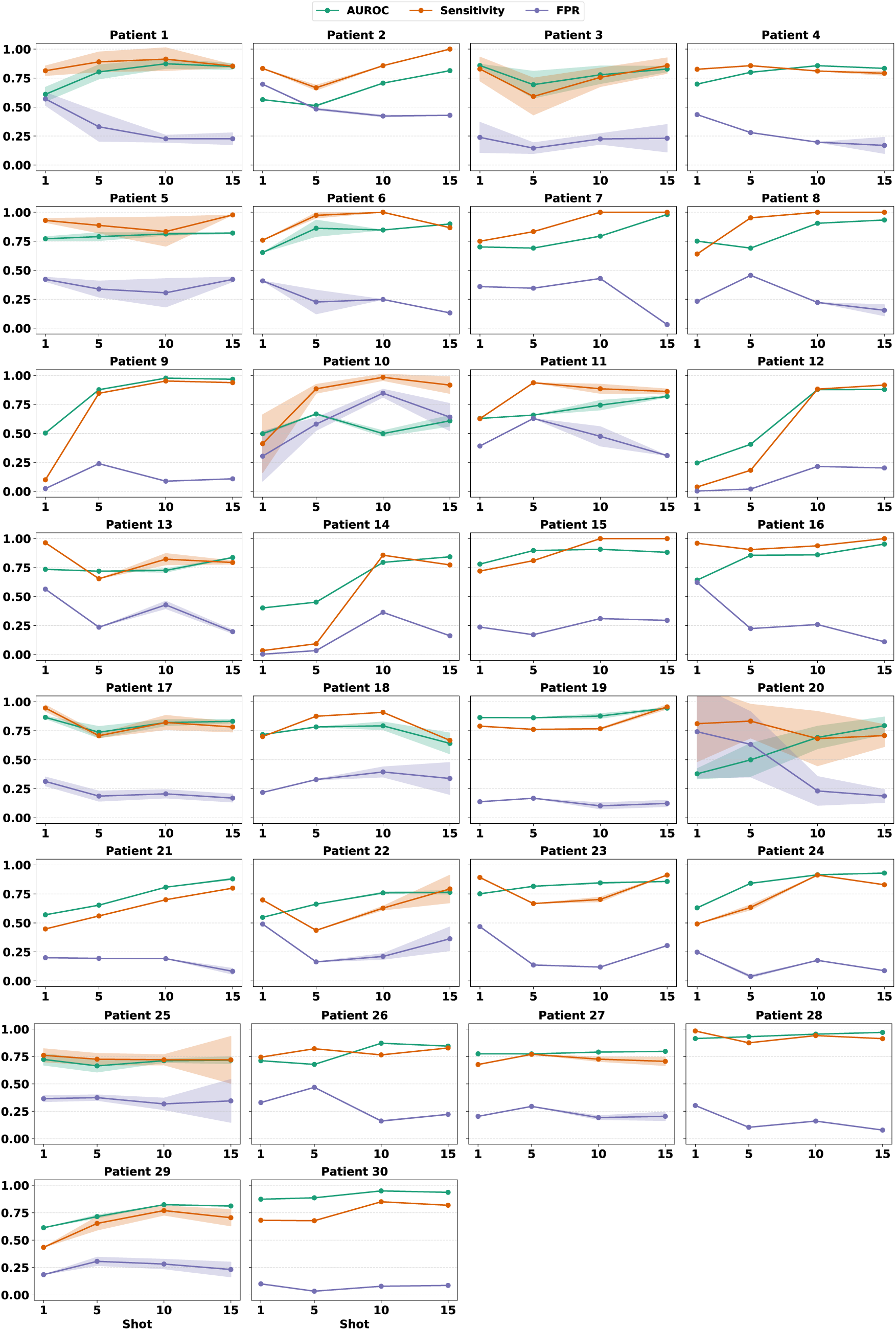
Performance metrics influenced as a function of shots for each patient in the EPILEPSIAE dataset. All results are averaged over 3 independent runs.

**Figure S3:**
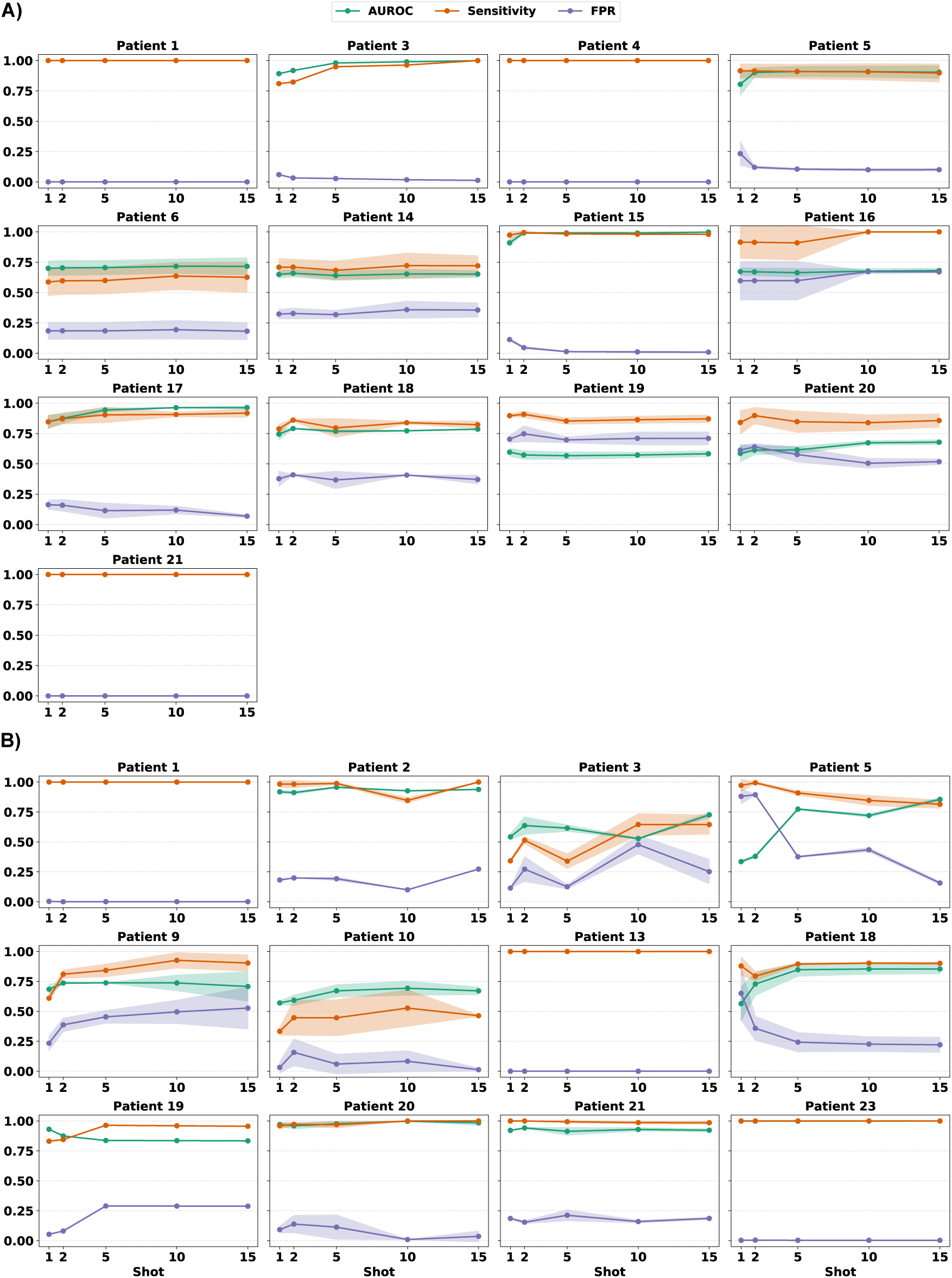
Performance metrics in the prediction framework as a function of shots for each patient in the FB (A) and CHB-MIT (B) datasets. All results are averaged over 3 independent runs.

### Supplementary Tables

**Table S1:** Comparison of CNN and our work for seizure detection on the EPILEPSIAE dataset across all patients.

| Patient | No. of Seizures | Interictal Hours | CNN [1] |  |  | OUR WORK |  |  |
| --- | --- | --- | --- | --- | --- | --- | --- | --- |
|  |  |  | AUROC | Sens | FPR | AUROC | Sens | FPR |
| Pat1 | 11 | 164.5 | **91.4** | 87.9 | **0.16** | 85.0 | **92.0** | 0.22 |
| Pat2 | 8 | 177.3 | 67.4 | 87.1 | 0.58 | **81.4** | **100.0** | **0.41** |
| Pat3 | 8 | 143.2 | **89.6** | **94.4** | 0.28 | 82.8 | 85.8 | **0.23** |
| Pat4 | 5 | 167.6 | **86.5** | 76.6 | 0.15 | 83.4 | **79.1** | **0.13** |
| Pat5 | 8 | 266.2 | 81.4 | 63.8 | **0.15** | **82.1** | **97.7** | 0.42 |
| Pat6 | 8 | 135.3 | **93.4** | 86.7 | **0.10** | 89.9 | **86.7** | 0.13 |
| Pat7 | 5 | 118.0 | 83.3 | 88.2 | 0.32 | **98.1** | **100.0** | **0.03** |
| Pat8 | 22 | 115.5 | 83.5 | 96.2 | 0.31 | **93.3** | **100.0** | **0.13** |
| Pat9 | 6 | 93.9 | 96.7 | 90.3 | **0.08** | **96.7** | **93.8** | 0.11 |
| Pat10 | 11 | 137.9 | 58.7 | 48.4 | **0.27** | **60.9** | **95.8** | 0.55 |
| Pat11 | 14 | 137.9 | **85.8** | 85.3 | **0.26** | 81.9 | **86.8** | 0.29 |
| Pat12 | 9 | 159.6 | **92.1** | **96.3** | 0.26 | 87.9 | 91.7 | **0.17** |
| Pat13 | 8 | 157.9 | 66.8 | **91.2** | 0.61 | **83.7** | 79.4 | **0.17** |
| Pat14 | 6 | 162.0 | **84.6** | **81.4** | 0.22 | 84.3 | 77.3 | **0.16** |
| Pat15 | 5 | 118.6 | **90.5** | 80.8 | **0.13** | 88.2 | **100.0** | 0.29 |
| Pat16 | 6 | 92.8 | 90.6 | 92.3 | 0.15 | **95.3** | **100.0** | **0.11** |
| Pat17 | 9 | 159.0 | **85.2** | **94.7** | 0.37 | 83.2 | 84.1 | **0.13** |
| Pat18 | 7 | 178.1 | **71.9** | **76.2** | **0.33** | 64.1 | 66.7 | 0.34 |
| Pat19 | 22 | 160.7 | 93.7 | 86.8 | 0.10 | **94.5** | **96.7** | **0.10** |
| Pat20 | 7 | 164.5 | **91.2** | **100** | 0.29 | 79.4 | 70.8 | **0.19** |
| Pat21 | 8 | 159.3 | 74.5 | **83.3** | 0.41 | **88.0** | 80.0 | **0.08** |
| Pat22 | 7 | 137.7 | 75.0 | 75.0 | 0.30 | **76.4** | **79.3** | **0.30** |
| Pat23 | 9 | 237.4 | **93.3** | 89.5 | **0.16** | 85.8 | **91.3** | 0.30 |
| Pat24 | 10 | 94.2 | 82.3 | 64.3 | 0.09 | **93.0** | **82.9** | **0.07** |
| Pat25 | 8 | 159.0 | 55.5 | **81.5** | 0.65 | **71.7** | 77.3 | **0.20** |
| Pat26 | 9 | 158.8 | **84.8** | **84.1** | **0.21** | 84.5 | 82.8 | 0.22 |
| Pat27 | 9 | 159.3 | 73.5 | **74.2** | 0.38 | **79.7** | 73.9 | **0.19** |
| Pat28 | 9 | 162.1 | 95.2 | 83.6 | **0.07** | **97.0** | **91.3** | 0.08 |
| Pat29 | 10 | 160.6 | 80.5 | 69.2 | 0.23 | **81.1** | **75.1** | **0.19** |
| Pat30 | 12 | 159.5 | **96.1** | **94.3** | 0.17 | 93.6 | 81.8 | **0.09** |
| **Total** | 276 | 4598.2 | 83.2 | 83.5 | 0.26 | **84.9** | **86.7** | **0.20** |

**Table S2:** Comparison of CNN, Future Guided Learning (FGL), and Our Work on CHB-MIT dataset across all patients.

| Patient | No. of Seizures | Interictal Hours | CNN [1] |  |  | FGL [2] |  |  | OUR WORK |  |  |
| --- | --- | --- | --- | --- | --- | --- | --- | --- | --- | --- | --- |
|  |  |  | AUROC | Sens | FPR | AUROC | Sens | FPR | AUROC | Sens | FPR |
| Pat1 | 7 | 17 | 92 | 85.7 | 0.24 | 98 | 89 | 0.04 | **100** | **100** | **0.00** |
| Pat2 | 3 | 22.9 | 34 | 33.3 | **0.00** | 88 | 83 | 0.10 | **93.8** | **100** | 0.09 |
| Pat3 | 6 | 21.9 | 95 | **100** | 0.18 | **97** | 90 | **0.01** | 72.6 | 64.5 | 0.46 |
| Pat5 | 5 | 13 | 88 | 80 | 0.19 | **96** | **97** | **0.07** | 85.5 | 81.4 | 0.16 |
| Pat9 | 4 | 12.3 | **74** | 50 | **0.12** | 57 | **94** | 0.60 | 70.8 | 90.4 | 0.48 |
| Pat10 | 6 | 11.1 | 56 | 33.3 | **0.00** | **81** | **65** | 0.26 | 67.1 | 46.4 | 0.22 |
| Pat13 | 5 | 14 | 95 | 80 | 0.14 | 100 | 99 | 0.00 | **100** | **100** | **0.00** |
| Pat18 | 6 | 23 | **93** | **100** | 0.28 | 69 | 88 | 0.43 | 85.4 | 90.1 | **0.27** |
| Pat19 | 3 | 24.9 | **98** | **100** | **0.00** | 76 | 89 | 0.48 | 83.4 | 95.6 | 0.09 |
| Pat20 | 5 | 20 | 97 | 100 | 0.25 | 94 | 86 | 0.13 | **98.5** | **100** | **0.01** |
| Pat21 | 4 | 20.9 | 89 | **100** | 0.23 | 90 | 73 | **0.12** | **92.2** | 98.5 | 0.15 |
| Pat23 | 5 | 3 | 99 | 100 | 0.33 | 67 | 100 | 0.50 | **100** | **100** | **0.00** |
| **Total** | 64 | 209 | 84.2 | 80.19 | 0.16 | 84.4 | 87.8 | 0.23 | **87.4** | **88.9** | **0.16** |

**Table S3:** Comparison of CNN and our Work for Seizure Prediction on the FB Dataset across all patients.

| Patient | No. of Seizures | Interictal Hours | CNN [1] |  |  | OUR WORK |  |  |
| --- | --- | --- | --- | --- | --- | --- | --- | --- |
|  |  |  | AUROC | Sens | FPR | AUROC | Sens | FPR |
| Pat1 | 4 | 23.9 | 100 | 100 | 0.00 | **100** | **100** | **0.00** |
| Pat3 | 5 | 23.9 | 100 | 100 | 0.00 | **100** | **100** | **0.00** |
| Pat4 | 5 | 23.9 | 100 | 100 | 0.00 | **100** | **100** | **0.00** |
| Pat5 | 5 | 23.9 | 65 | 40 | 0.13 | **90.4** | **89.7** | **0.10** |
| Pat6 | 3 | 23.8 | **100** | **100** | **0.00** | 71.6 | 62.6 | 0.18 |
| Pat14 | 4 | 22.6 | **82** | 50 | **0.27** | 65.2 | **72.1** | 0.36 |
| Pat15 | 4 | 23.7 | 99 | **100** | 0.02 | **99.8** | 98.1 | **0.00** |
| Pat16 | 5 | 23.9 | **86** | 80 | **0.17** | 68.1 | **100** | 0.67 |
| Pat17 | 5 | 24.0 | 94 | 80 | **0.00** | **96.4** | **91.8** | 0.07 |
| Pat18 | 5 | 24.8 | **100** | **100** | **0.00** | 78.6 | 83.7 | 0.37 |
| Pat19 | 4 | 24.3 | 51 | 50 | **0.16** | **58.3** | **87.1** | 0.71 |
| Pat20 | 5 | 24.8 | **76** | 60 | **0.04** | 67.9 | **85.7** | 0.52 |
| Pat21 | 5 | 23.9 | 100 | 100 | 0.00 | **100** | **100** | **0.00** |
| **Total** | 59 | 311.4 | **88.7** | 81.54 | **0.06** | 84.3 | **90.1** | 0.23 |

